# Mortality and vaccine-preventable deaths from severe acute respiratory infections in Brazil in 2026

**DOI:** 10.64898/2026.10.03.26364639

**Authors:** Emil Kupek

## Abstract

**Background:** A large increase in mortality from severe acute respiratory infections (SARI) in Brazil in 2025 was mainly driven by influenza A and respiratory syncytial virus (RSV) among children and the aged, but 2026 data remain largely unpublished. Methods – Secondary data on SARI by age group (<2, 2-4, 5-14, 15-49, 50-64, 65+ years) and virus (Influenza A/B, RSV, SARS-CoV-2, adenovirus, rhinovirus, bocavirus, and metapneumovirus) were retrieved for 35 weeks of 2026. Weekly variations in infection rate (IR) and mortality rate (MR) were described by virus and age group. Case fatality ratio (CFR) and vaccine-preventable deaths from influenza, RSV, and SARS-CoV-2 were analysed by age group. Missing virus classification was estimated by multinomial regression. Poisson-based 95% confidence intervals (CI) were calculated for MR.

**Results:** Influenza A infections quintupled during February and March, with a mortality peak in April, mainly due to H2N3. RSV and rhinovirus caused more deaths than influenza viruses, especially among young children (<2) and the aged (65+). The former were most frequently infected by RSV and the latter by rhinovirus. For those 50 years or older, influenza B reached a CFR of about 20%, compared with 5-15% for influenza A, metapneumovirus, rhinovirus, and other non-subtyped viruses. The highest vaccine-preventable MRs per 100,000 were 17.23 for the oldest and 8.62 for the youngest. Under ideal conditions, 6418 lives could have been saved, about half among children and adolescents <15 years old.

**Conclusions:** Vaccine-preventable SARI in Brazil exerted a high toll in fall and winter 2026, mainly due to influenza A, RSV, and rhinovirus. Better vaccine coverage may significantly reduce this toll.

## 1. Introduction

Severe acute respiratory infections (SARI) remain a major cause of hospital admissions and mortality worldwide, particularly in low-and middle-income countries [1], despite a sharp decline in the SARS-CoV-2 share of this burden. Brazil is no exception: hospital admissions and mortality rates (MRs) related to these infections dropped significantly between 2020 and 2025 [2]. However, there was a large increase in these rates in 2025 [3], driven primarily by atypical influenza A [4] and respiratory syncytial virus (RSV).

SARI surveillance in Brazil is maintained by the Ministry of Health’s epidemiological surveillance [5], updated daily with medical records from the federal states, verified for inconsistencies, and published in an abbreviated version on the Internet [6]. SARI bulletins by Fiocruz [7] synthesise key trends and alert to imminent risk by geographic region.

However, underestimation of cases and deaths is frequent with routinely collected epidemiological surveillance data in Brazil due to reporting delays, the lack of laboratory data, and limited access to medical care in Brazil. The COVID-19 pandemic prompted improvements in the timeliness and completeness of surveillance data. For example, COVID-19 data entry during 2020-2023 decreased from about two to one week, and 86% of suspected cases received a final diagnosis [8]. Nevertheless, another study reported 37.2%, 22.0%, 44.8%, and 56.4% of SARI without an etiological agent for successive years of the same period [9].

Although SARS-CoV-2 is no longer a predominant cause of SARI and its vaccine prevention has been available at no cost in the public sector, COVID-19 has kept a significant share of the SARI burden in Brazil. It caused almost 23% of all SARI deaths whose pathogen was identified in 2025 [10]. Other vaccines routinely applied in the country to prevent respiratory infections are against influenza and respiratory syncytial virus (RSV), both of which have been leading causes of SARI since the decrease [2,3]. In Brazil, RSV-related hospital admissions increased sharply during and after the COVID-19 pandemic and passed 15 per 100,000 in 2024, mainly in southern and southeastern regions, which also have better laboratory testing capacity [11]. Case fatality ratio (CFR) ranged between 21% and 31%, while in-hospital mortality increased fivefold over the five years analysed, with cardiovascular diseases and diabetes as the most prevalent risk factors [11].

In 2026, SARI bulletins [7] and news agencies alerted that influenza A and RSV were leading causes of hospital admissions and SARI mortality in April 2026, with children under 2 years and the aged most susceptible [12]. By the end of April, over 46,000 cases and almost 2,000 deaths were attributed to SARI, with rhinovirus, influenza A, RSV, and SARS-CoV-2 as leading causes of infection, whereas influenza A, SARS-CoV-2, and rhinovirus were leading causes of death. The latter sequence is similar to the 2025 ranking of major causes of SARI mortality [10]. In June, all age groups showed an increase in SARI infections and related mortality, but those at the extremes of age distribution were most affected. Influenza A, rhinovirus, and RSV continued to be the key drivers of this trend, with the former responsible for almost half of deaths [10,13]. Rhinovirus and RSV had CFR in the range of 16-17%, followed by SARS-CoV-2 with a CFR of 9%.

By the second week of September, SARI incidence and mortality rates declined, with a shift in the most affected age group to 2-14-year-olds [14]. In children <2 years, RSV, rhinovirus, and metapneumovirus were the main causes of SARI-related deaths. Between January and mid-September, leading causes of SARI mortality were influenza A (35.4%), rhinovirus (22.9%), RSV (16,2%), SARS-CoV-2 (14%), and influenza B (9.3%).

In addition to RSV and influenza, laboratory kits used for routine epidemiological surveillance in Brazil also detect adenovirus, bocavirus, metapneumovirus, and rhinovirus. The specific contributions of each of these viruses to SARI mortality were described for the pre-COVID-19 period [15], but have been sparsely reported since. The present study aims to provide the first estimates of infection and mortality rates for key SARI viruses in Brazil in 2026 and account for missing virus identification.

## 2. Methods

This study was conducted with secondary data on SARI infections and deaths in Brazil during the first 35 weeks of 2026. Brazilian Ministry of Health maintains two sites with daily updates on SARI: SIVEP-Gripe and e-SUS. The latter includes mild and moderate cases of COVID-19, whereas the former covers all SARI-related deaths and infections caused by influenza, RSV, SARS-CoV-2, adenovirus, bocavirus, metapneumovirus, rhinovirus, and other unidentified respiratory viruses. The H1N1 and H3N2 subtypes are available for influenza A. Both sources are unified, curated [16], and kept in the public domain [6].

Laboratory results were originally classified as confirmed, inconclusive, and absent. The last two were considered missing for this study.

Multinomial regression was used to impute the unidentified causative agents of SARI based on the following independent variables: federal state, age group, week of the year, and its quadratic and cubic terms. Missing values were substituted with predicted probabilities derived from the regression based on complete data. Robust standard errors were calculated because the data were clustered within municipalities of patient residence. Epidemiological surveillance is part of municipal administration, so eventual delays and misdiagnoses would originate from the same source and therefore induce correlations between reports.

Age groups included <2, 2-4, 5-14, 15-49, 50-64, and 65 years or older. Population size for these groups was obtained from official population statistics [17].

The numbers of SARI infections and deaths were divided by population size within age groups to calculate infection (IR) and mortality (MR) rates. Poisson-based 95% confidence interval (CI) was used to express uncertainty of mortality rates, whereas a binomial distribution was assumed for proportion/percentage statistics.

The number of vaccine-preventable deaths was calculated by multiplying mortality rates by population size for influenza, COVID-19, and RSV. These vaccines are available in Brazilian public health facilities at no cost to users, but the RSV vaccine is limited to specific at-risk groups. The percentage of vaccine-preventable deaths was calculated as the number of these deaths divided by total SARI deaths, multiplied by 100.

Version 17.0 of Stata software was used for data management, graphs, and statistics.

## 3. Results

Multinomial regression estimated that most of the 14,652 SARI cases without virus identification (38.48% of total) were likely rhinovirus (38.9%), influenza A (29.7%), RSV (25.27%), COVID-19 (3.62%), and influenza B (2.36%).

Influenza A infection rate quintupled between February and March, while related mortality doubled between the beginning of March and the end of April, with A(H2N3) as the key component (Figure 1). Influenza B infections and related mortality increased during May and June, with magnitude similar to that of influenza A(H2N3).

**Figure 1.**
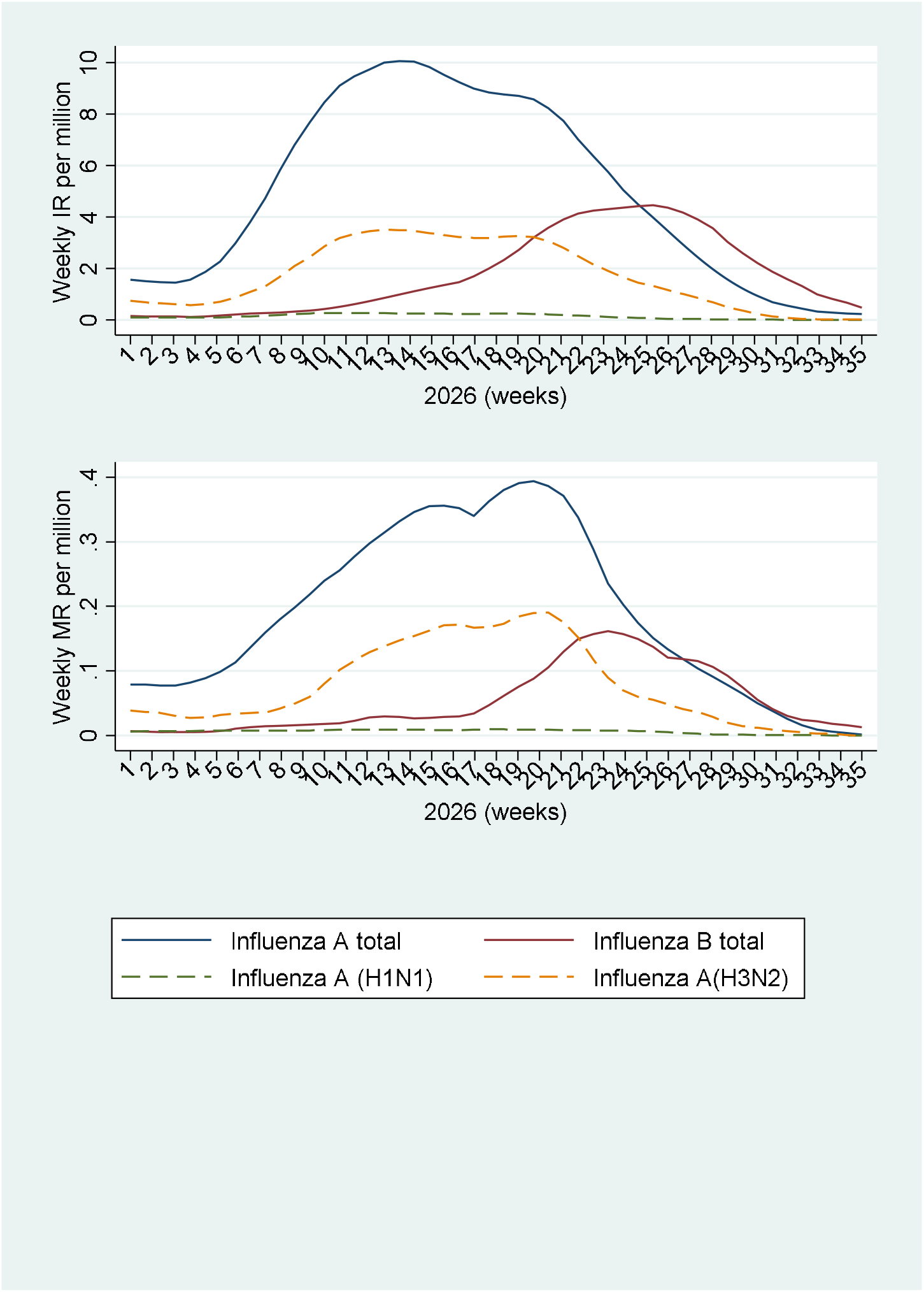
Mortality (MR) and infection (IR) rates for influenza viruses causing SARI.

RSV and rhinovirus together caused more infections and deaths than influenza viruses (Figure 2). SARS-CoV-2 maintained a low IR with decreasing mortality. Adenovirus, metapneumovirus, and other respiratory viruses kept low-level IR and MR over the period analysed.

**Figure 2.**
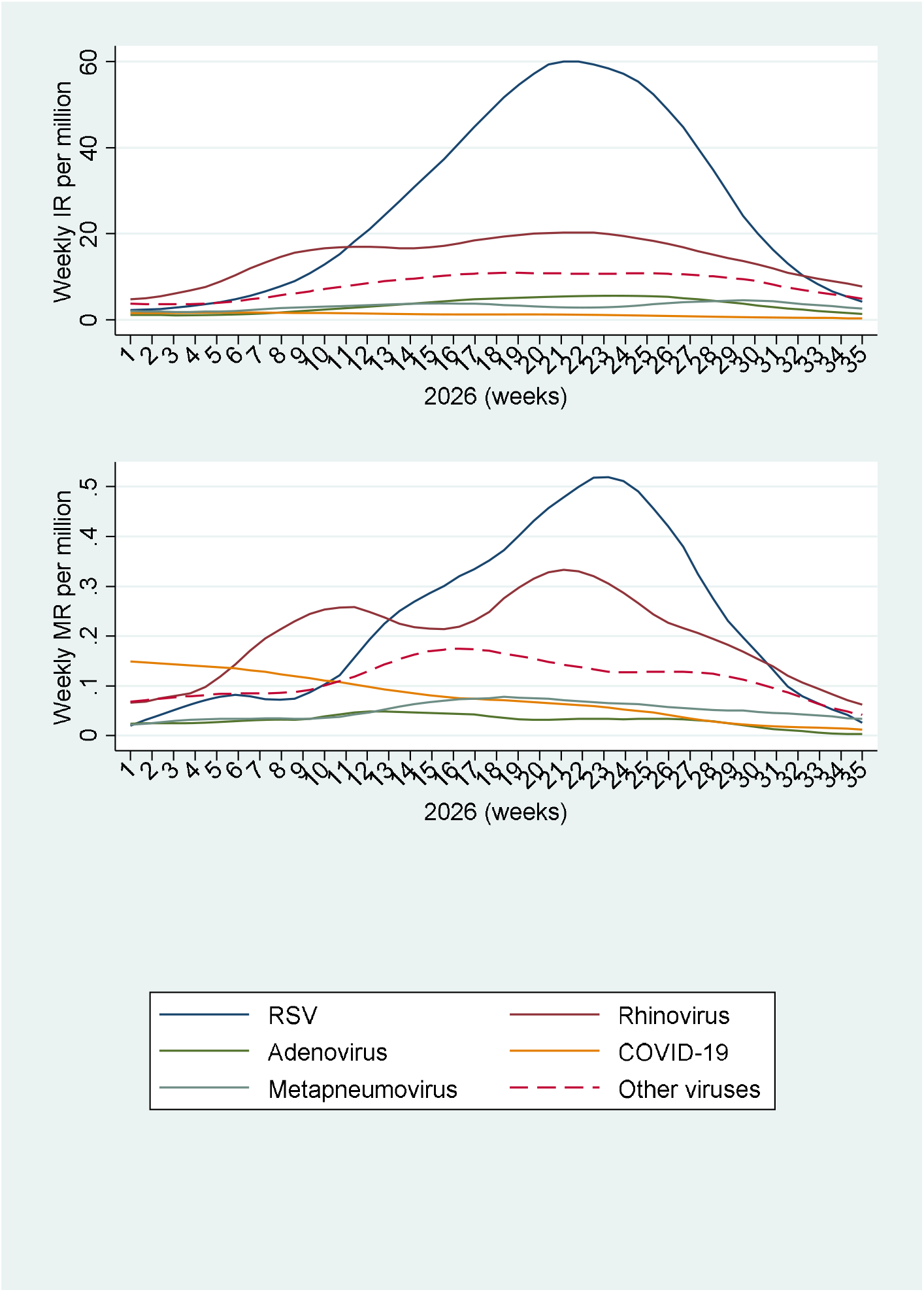
Mortality (MR) and infection (IR) rates for non-influenza viruses causing SARI.

Influenza mortality was highest among the aged (65+ years) and children <2 years (Figure 3). The latter showed increasing influenza A mortality between February and March, reached a plateau in April, and initiated its decline in May. Influenza B-related mortality rose steeply in April and May, then fell rapidly. Among the oldest age group analysed, influenza A mortality began to rise in February, peaked by the end of April, then decreased to almost zero by mid-September. Influenza B showed a similar seasonal pattern to influenza A, but at about three times lower magnitude.

**Figure 3.**
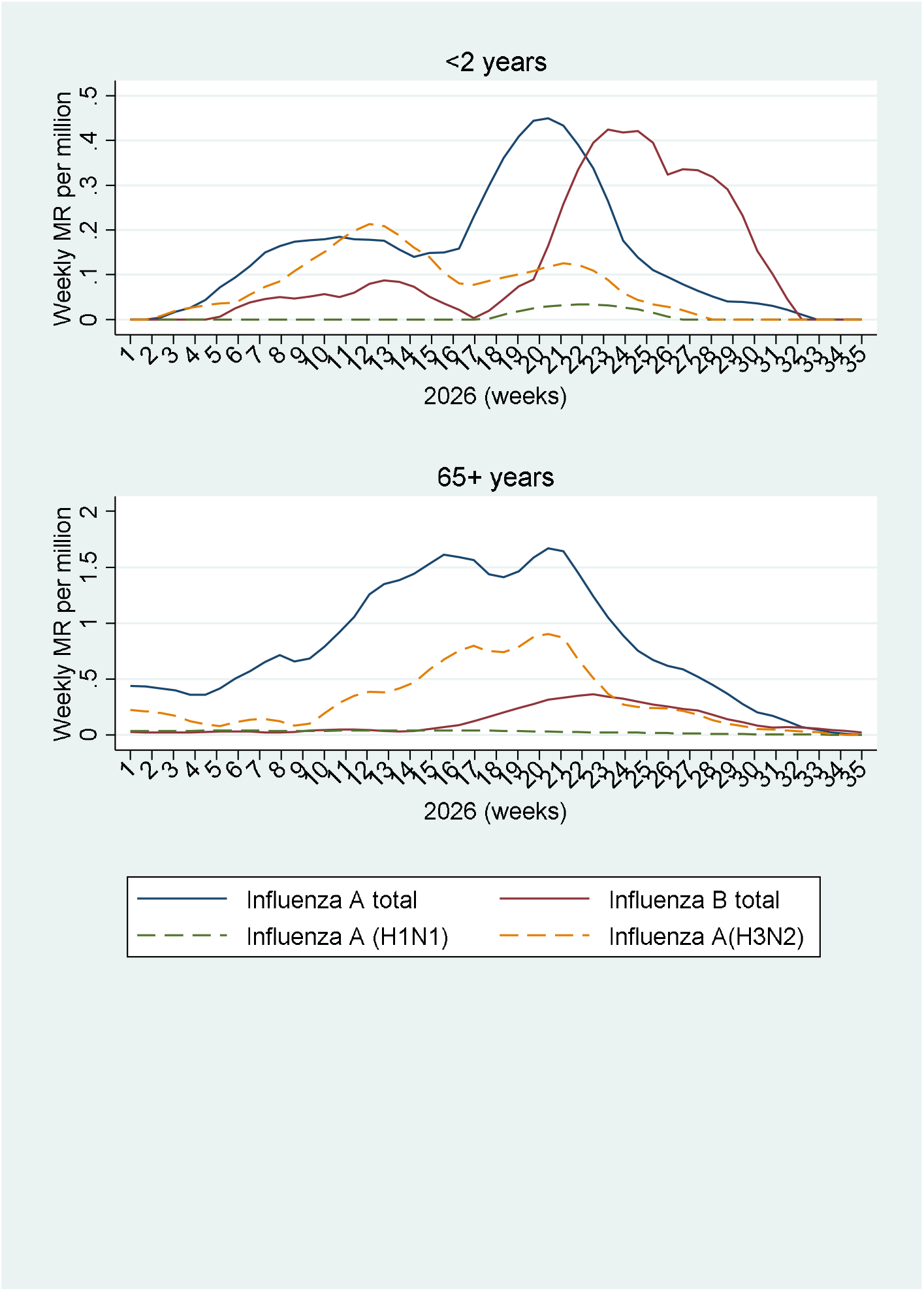
Mortality rate (MR) by influenza viruses causing SARI for children <2 years and the aged (65+ years)

Among non-influenza viruses causing SARI, RSV was the most salient for children <2 years and rhinovirus for the aged (Figure 4). The latter also registered declining COVID-19 mortality over the period analysed and higher mortality from other viruses.

**Figure 4.**
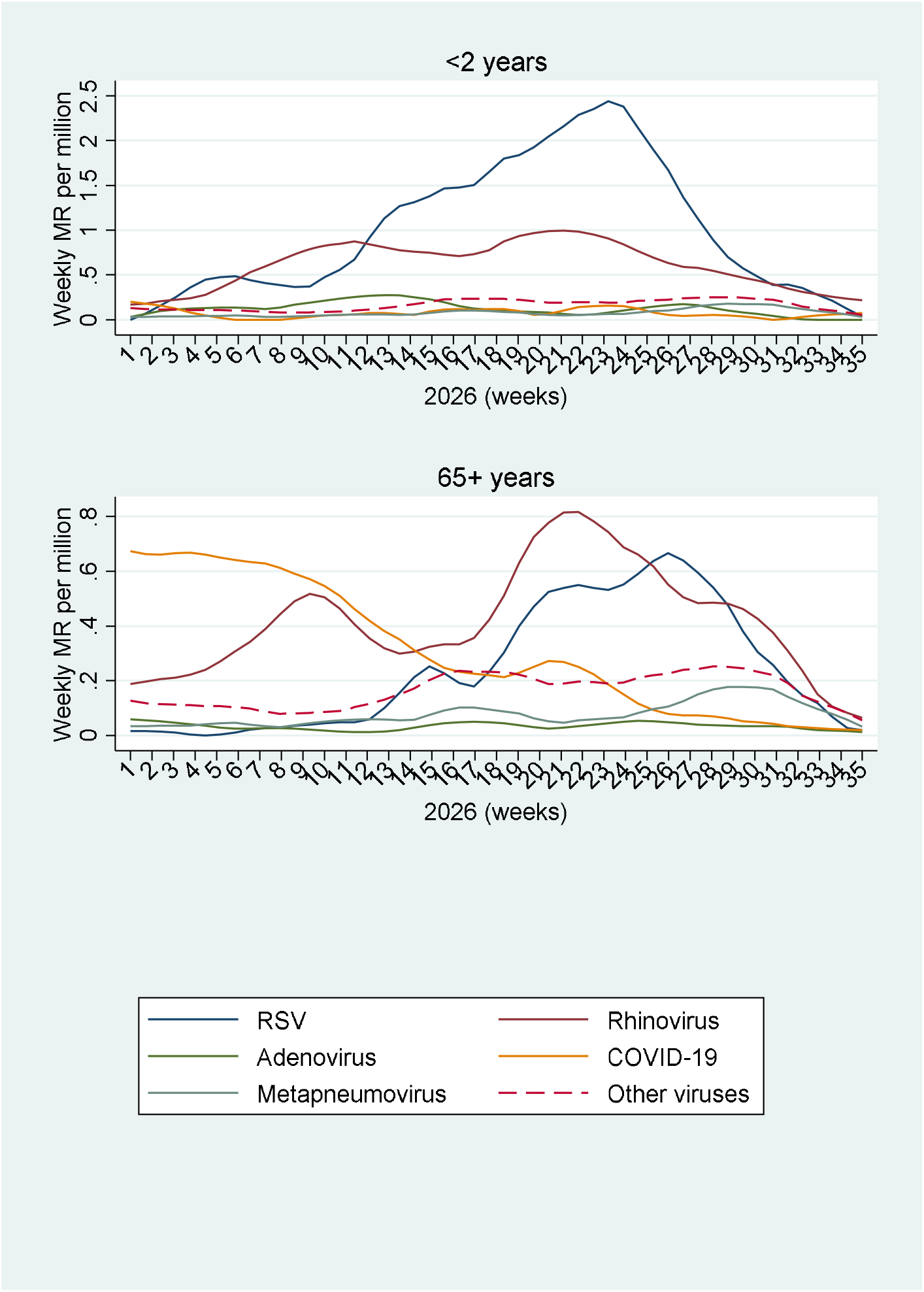
Mortality rate (MR) by non-influenza viruses causing SARI for children <2 years and the aged (65+ years)

Case fatality often showed a rising gradient across age groups for most viruses (Table 1). COVID-19 was an exception, with a CFR of almost 2% for the youngest and oldest groups. In children <2 years, CFR for influenza B was 1.77%, about twice that of influenza A, while CFRs for metapneumovirus, rhinovirus, and other non-subtyped respiratory viruses were close to 1%. In adults aged 15-49 years, CFRs for influenza B, metapneumovirus, adenovirus, and other non-subtyped viruses fall in the range of 8-10%, and for influenza A, RSV, and rhinovirus in the range of 4-8%. For those 50 years or older, influenza B reached a CFR of about 20%, while CFRs for influenza A, metapneumovirus, rhinovirus, and other non-subtyped viruses ranged between 5% and 15%.

**Table 1.** Number of cases, deaths, and case fatality ratio (CFR) for respiratory viruses in Brazil, 2026 (up to the 35^th^ week)

| Virus | Outcome | Age (years) |  |  |  |  |  |
| --- | --- | --- | --- | --- | --- | --- | --- |
|  |  | <2 | 2-4 | 5-14 | 15-49 | 5-64 | 65+ |
| Adenovirus | cases | 3094 | 808 | 287 | 94 | 52 | 108 |
|  | deaths | 23 | 4 | 3 | 9 | 4 | 29 |
|  | CFR (%) | 0.73 | 0.53 | 1.09 | 9.76 | 7.84 | 2.71 |
| Bocavirus | cases | 419 | 89 | 42 | 8 | 12 | 14 |
|  | deaths | 3 | 0 | 1 | 3 | 3 | 2 |
|  | CFR (%) | 0.73 | 0 | 2.39 | 3.76 | 25.00 | 1.43 |
| COVID-19 | cases | 856 | 149 | 132 | 653 | 883 | 2848 |
|  | deaths | 17 | 2 | 8 | 73 | 161 | 566 |
|  | CFR (%) | 1.98 | 1.37 | 6.23 | 1.12 | 1.83 | 1.99 |
| Influenza A | cases | 3188 | 1609 | 2381 | 7405 | 5921 | 20085 |
|  | deaths | 30 | 17 | 31 | 428 | 634 | 2688 |
|  | CFR (%) | 0.95 | 1.05 | 1.31 | 5.77 | 10.70 | 13.38 |
| Influenza B | cases | 1277 | 439 | 1161 | 2294 | 435 | 674 |
|  | deaths | 23 | 4 | 23 | 217 | 84 | 145 |
|  | CFR (%) | 1.77 | 0.99 | 1.95 | 9.44 | 19.26 | 21.56 |
| Influenza, no subtype | cases | 137 | 59 | 78 | 65 | 38 | 158 |
|  | deaths | 3 | 0 | 1 | 8 | 5 | 38 |
|  | CFR (%) | 2.27 | 0 | 1.32 | 11.75 | 13.56 | 24.12 |
| Metapneumovirus | cases | 3047 | 594 | 248 | 152 | 174 | 537 |
|  | deaths | 34 | 6 | 4 | 13 | 17 | 70 |
|  | CFR (%) | 1.10 | 1.06 | 1.69 | 8.26 | 9.72 | 13.13 |
| Rhinovirus | cases | 19786 | 11058 | 16991 | 4300 | 3688 | 7529 |
|  | deaths | 188 | 46 | 84 | 183 | 214 | 738 |
|  | CFR (%) | 0.95 | 0.41 | 0.49 | 4.25 | 5.81 | 9.80 |
| RSV | cases | 62362 | 9387 | 1391 | 496 | 903 | 7081 |
|  | deaths | 361 | 45 | 7 | 38 | 73 | 806 |
|  | CFR | 0.58 | 0.48 | 0.51 | 7.72 | 8.08 | 111.38 |
| Other<br>respiratory<br>virus | cases | 8073 | 1867 | 836 | 418 | 354 | 1001 |
|  | deaths | 86 | 15 | 10 | 38 | 37 | 152 |
|  | CFR (%) | 1.07 | 0.79 | 1.20 | 9.09 | 10.52 | 15.22 |
| Total | cases | 102238 | 26059 | 23549 | 15885 | 12459 | 40035 |
|  | deaths | 767 | 139 | 172 | 1009 | 1232 | 5235 |
|  | CFR | 0.75 | 0.53 | 0.73 | 6.35 | 9.89 | 13.08 |
| Vaccine-preventable<br>(Influenza,<br>RSV,<br>Covid-19) | deaths | 434 | 68 | 70 | 764 | 957 | 4243 |
|  | Pop.<sup>a</sup> | 5.0 | 7.7 | 28.5 | 112.7 | 36.1 | 24.4 |
|  | MR<sup>b</sup> | 8.68 | 0.88 | 0.25 | 0.68 | 2.65 | 17.39 |
|  | (95% CI) | (7.88, 9.54) | (0.68, 1.11) | (0.19, 0.31) | (0.63, 0.73) | (2.49, 2.82) | (16.87, 17.92) |
|  | PVPD<sup>c</sup> | 56.58<br>(52.99, 60.12) | 48.92<br>(40.35, 57.53) | 40.70<br>(33.28, 48.43) | 75.72<br>(72.95, 78.33) | 77.68<br>(75.24, 79.98) | 81.05<br>(79.96, 82.10) |

<sup>a</sup> Population in millions
<sup>b</sup> Mortality rate per 100,000 (95% confidence interval)
<sup>c</sup> Percentage of vaccine-preventable deaths: (vaccine-preventable deaths / total deaths) x 100

SARS-CoV-2, influenza, and RSV are all vaccine-preventable. Ideally, with 100% vaccine coverage and effectiveness, we would prevent all deaths from these viruses. Although this is not realistic, it serves as a benchmark and upper bound for vaccine effectiveness. Vaccine-preventable MRs per 100,000 by age group were 17.23 for the oldest, 8.62 for the youngest, 2.64 for 50-64-year-olds, and <1 for other age groups (Table 1). These rates amount to 6,418 lives potentially saved, including 431 children <2 years, 755 15-49-year-olds, 953 50-64-year-olds, and 4,204 people of 65 years or older. Relative to total SARI deaths, we would save 40-78% of lives lost to SARI in each age group. Of note, this includes 40-57% of children and adolescents under 15 years.

## 4. Discussion

To the best of my knowledge, this study is the first comprehensive analysis of SARI-related mortality in Brazil in 2026 by virus type and age, with imputation of missing virus identification to avoid underestimation. The latter was close to 40% and highlights the downward bias of estimates based only on confirmed cases. Another study pointed out that about 35% of SARI cases and 18% of related deaths did not specify the cause of infection in the 2020-2025 period [2]. As COVID-19 dominance waned, mortality attributed to “other viruses” increased [2]. In 2025, just over half of notified SARI cases had pathogen identification [10].

The results reveal that influenza A, particularly its H3N2 strain, contributed most to overall influenza mortality in 2026. Influenza B mortality peaked in June, about a month after the influenza A peak. Influenza A(H3N2) was the most salient component of the 2025 season, when a drifted clade initiated the second wave of influenza in August [4].

In the present study, people 65 years or older and young children (<2 years) registered the highest influenza mortality, mainly due to A(H3N2), whereas influenza B ranked high among young children. Influenza B peaked earlier in these two most vulnerable groups compared to other age groups.

Among the children under two years, RSV was the foremost contributor to SARI mortality, with rhinovirus in the second place. In the oldest group, however, rhinovirus had the largest share, and RSV ranked second among causes of SARI mortality. The overall RSV-related mortality was similar to influenza A mortality, followed by rhinovirus and non-subtyped viruses.

CFR was highest in adults and those aged 50 years or older, with influenza B at about 20%, and influenza A, metapneumovirus, and RSV in the range of 8-15%. High case fatality is likely due to comorbidities often found in older age groups, as highlighted in another Brazilian study which covered the 2013-2024 period [11]. The latter also found CFRs in the range of 21-31% in people with 60 years or older. However, the present study found that the 50-64-year-olds had CFRs almost as high as the oldest age group.

Among study limitations, reporting delay and misdiagnosis are probably the most important sources of bias. The former is certainly present in September data and, to a lesser extent, the August data, both of which are likely underreported. Nevertheless, the study results are still useful as a lower bound for SARI mortality. Missing virus identification was addressed by imputation, whose accuracy depends on the quality of the data used to estimate missing diagnoses and the assumed relationship between observed and missing data. Although uncertainty of MR is certainly underestimated, mean values are likely contained within the confidence intervals. Furthermore, without whole-year data available in mid-September, seasonality analysis remains limited, especially for the end-of-winter period in Brazil. Despite these limitations, the study estimates are less biased than those based exclusively on confirmed viral identification, which disregards a large portion of the data.

Further research should update SARI records to provide better estimates of seasonal variation for the most common respiratory viral diseases and improve the precision of these estimates. Egg-based influenza vaccines need about six months to scale up production, so to start mass vaccination in March next year, data up to September provide critical input on circulating strains, whose virulence and pathogenicity in the following season can be gauged only by partial data from the previous year.

Brazilian SARI surveillance critically depends on the capacity of local health authorities to gather and process health records in real time. Delayed laboratory results and administrative constraints lead to underestimation of SARI burden and its key components. However, timely analysis and regular use of statistical tools may improve understanding of these infections and increase preparedness to prevent them.

Vaccination remains the most powerful tool in preventing serious SARI-related diseases. The 52.4% influenza A vaccine coverage in the target population [4] was insufficient to prevent a large increase in mortality in 2025 [18], and 2026 SARI mortality appears to follow the same pattern. A large-scale RSV vaccination of pregnant women and timely treatment of at-risk young children with monoclonal antibodies (nirsevimab) may significantly reduce paediatric mortality [10].

## 5. Conclusions

Between January and mid-September 2026, SARI mortality in Brazil remained at a high level reported for the previous year, with similar age distribution and similar pathogens (influenza A, especially H3N2 subtype, RSV, rhinovirus) as leading causes of death. Young children and the aged were most vulnerable to these viruses. With 100% vaccination coverage and effectiveness, 6,418 lives lost to SARI would be saved. Under realistically achievable coverage and effectiveness, a significant number of SARI deaths would still be prevented.

## Data Availability

All data produced are available online at
https://gitlab.com/cgcovid/dados-publicos/blob/main/Dados%20unificados/README.md?ref_type=heads

## Notes

### Competing Interest Statement

The authors have declared no competing interest.

